# Inflammatory profiles in sputum and blood of people with TB with and without HIV coinfection

**DOI:** 10.1101/2024.04.23.24306127

**Authors:** Sara C Auld, Artur T L Queiroz, Mariana Araujo-Pereira, Pholo Maenetje, Nomsa Mofokeng, Lerato Mngomezulu, Duduzile Masilela, Brian Dobosh, Rabindra Tirouvanziam, Hardy Kornfeld, Bruno B Andrade, Gregory P Bisson

## Abstract

Although tuberculosis (TB) remains a major killer among infectious diseases and the leading cause of death for people with HIV, drivers of immunopathology, particularly at the site of infection in the lungs remain incompletely understood. To fill this gap, we compared cytokine profiles in paired plasma and sputum samples collected from adults with pulmonary TB with and without HIV. We found that people with pulmonary TB with HIV had significantly higher markers of inflammation in both plasma and sputum than those without HIV; these differences were present despite a similar extent of radiographic involvement. We also found that the strength and direction of correlations between biomarkers in the blood and lung compartments differed by HIV status and people with HIV had more positive correlations than those without HIV. Future studies can further explore these differences in inflammation by HIV status across the blood and lung compartments and seek to establish how these profiles may be associated with long-term outcomes and lung health after completion of TB treatment.

---

Tuberculosis (TB) remains a major killer among infectious diseases and the leading cause of death for people with HIV. TB-associated inflammation aids in pathogen killing but can lead to unwanted lung damage. Although therapeutic strategies to modify host immune responses could decrease tissue damage, improve treatment outcomes, and reduce post-TB morbidity, drivers of immunopathology remain incompletely understood.

HIV is associated with impaired pulmonary epithelial integrity, chronic pulmonary inflammation (1), and altered circulating cytokine profiles in people with TB (2). Yet, few studies have compared cytokine profiles in the lungs of TB patients with and without HIV. This is important as pulmonary immune responses to *Mycobacterium tuberculosis* (*Mtb*) may differ from those in blood (3-6). To fill this gap, we compared cytokine profiles in paired plasma and sputum samples collected from adults with pulmonary TB with and without HIV.

## Methods

We conducted a cross-sectional study in Johannesburg, South Africa during August 2021-April 2022. Adults with a first episode of Xpert MTB/RIF-positive pulmonary TB without rifampin resistance who had initiated treatment in the previous 8 weeks with at least with moderate or severe involvement on chest radiograph (CXR) were enrolled. Patients who were pregnant, incarcerated, had a diagnosis of COVID-19 within 90 days of screening, or had TB meningitis were excluded. HIV testing was performed for those with an unknown HIV status.

Participants completed a medical history questionnaire and, for those with HIV, a recent CD4 cell count was abstracted from local clinic records. Plasma and induced sputum were collected and stored at −80°C before shipment to the FIOCRUZ laboratory in Salvador, Brazil. Plasma and sputum supernatants were analyzed by bead-based multiplex assay with the Luminex™ xMAP™ INTELLIFLEX System (Milliplex MAP Human Cytokine/Chemokine Magnetic Bead Panel).

Descriptive statistics were stratified by HIV. For cytokine analyses, hierarchical clustering was used to display z-score normalized data using the ComplexHeatmap package (version 2.16.0). Log2-fold-changes were measured (gtools version 3.9.4) and median cytokine concentrations were compared with the Mann-Whitney *U* test. Correlation analysis was performed with the Spearman rank test, with significant correlations (p-value < 0.05) plotted with the circlize package (version 0.4.15). All analyses were performed in R 4.2.1.

The Institutional Review Board of Emory University and the Health Research Ethics Counsel in South Africa approved this study. All participants provided written informed consent.

## Results

Among 30 participants enrolled, the median age was 32 (interquartile range [IQR] 27-40), 6 (20%) were female, median body-mass index was 18.9 kg/m^2^ (IQR 17.0-20.1), and 16 (53%) reported ever smoking (Table 1). Ten participants had HIV coinfection, of whom 5 were on ART. The median CD4 count was 197 cells/μl (IQR 31-246). There were 12 (40%) participants with moderate and 18 (60%) with severe CXR involvement. There were no significant differences in demographic or clinical variables by HIV status.

**Table.** Demographic and clinical characteristics of study participants.

|  | **All (n = 30)** | **HIV- (n = 20)** | **HIV+ (n = 10)** | **p-value** |
| --- | --- | --- | --- | --- |
| Age (median, IQR) | 32 (27-40) | 33 (26 – 41) | 31 (28 – 37) | 0.69 |
| Female (n, %) | 6 (20%) | 3 (15%) | 3 (30%) | 0.33 |
| Ever smoker (n, %) | 16 (54%) | 11 (55%) | 5 (50%) | 0.79 |
| Cigarette pack-years (median, IQR) | 4.0 (3.0 – 7.6) | 4.0 (3.0 – 7.6) | 3 (3.0 – 7) | 0.42 |
| BMI (median, IQR) | 18.9 (17 – 20.1) | 18.6 (16.9 – 19.6) | 19.0 (18.0 – 21.2) | 0.50 |
| ART status (n, %) | -- | -- | 5 (50%) | n/a |
| CD4 count (median, IQR)* | -- | -- | 197(31 – 246) |  |
| Duration of cough (weeks; median, IQR) | 4 (4 – 6) | 4 (4 – 6) | 4 (4 – 6) | 0.77 |
| *CXR severity* |  |  |  |  |
| Moderate CXR (n, %) | 10 (33%) | 7 (35%) | 3 (30%) | 0.43 |
| Severe CXR (n, %) | 18 (60%) | 13 (65%) | 5 (50%) |  |

*\*CD4 count available for 9 participants*

Median sputum biomarker concentrations were higher for 24 of 29 cytokines among participants with HIV than those without HIV, and were significantly higher for IL-1α, EGF, and VEGF (p-value < 0.05) (Figure 1A). Similarly, median plasma concentrations for 21 of 29 cytokines measured were higher in those with HIV, and this difference was statistically significant for IL-1α, IL-1RA, IL-6, IL-10, IP-10/CXCL10, G-CSF, and TNF-α (Figure 1A). Cytokine concentrations were at least 2-fold higher in sputum than plasma for 11 of 29 cytokines.

**Figure.**
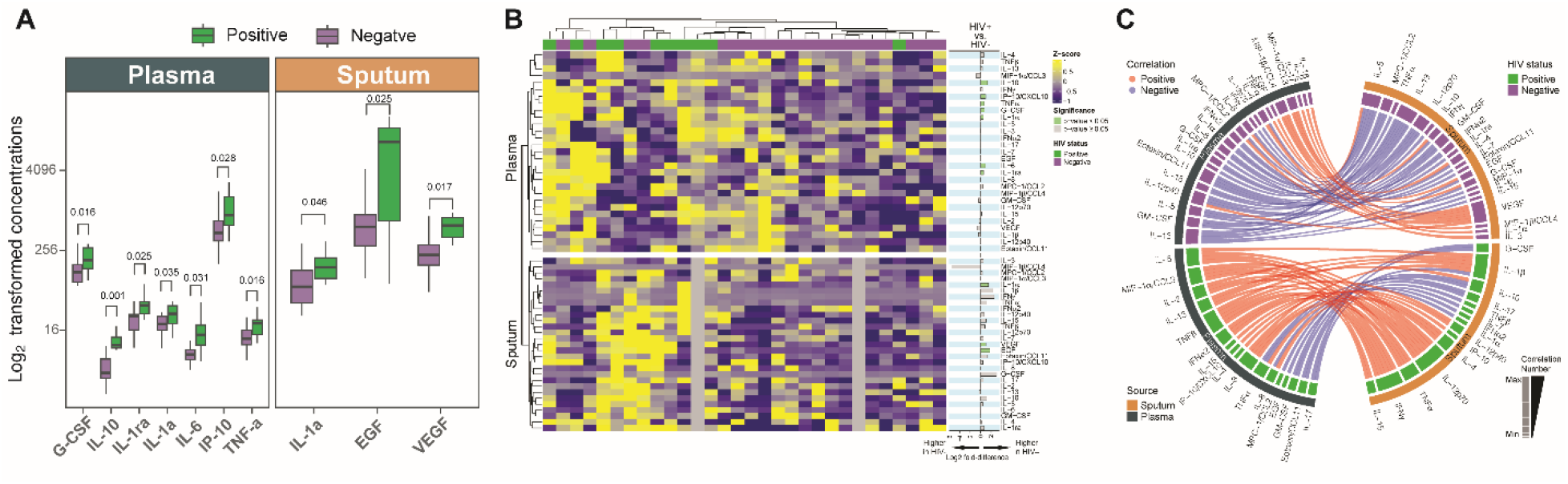
Log_2_-transformed cytokine concentrations (A), heatmap showing z-scores (B), and Circos plot with correlations (C) for sputum and plasma cytokines for people with pulmonary TB, with and without HIV. (A) Log2-transformed cytokine concentrations with statistically significant differences by HIV status for seven cytokines measured in plasma and three cytokines measured in sputum. (B) Heatmap with z-scores for plasma (top half) and sputum (bottom half) cytokine concentrations after hierarchical clustering. HIV status is shown across the top, with people with HIV in green and people without HIV in purple. Log2-fold-change in cytokine concentrations for those with vs. without HIV is shown on the right; purple shading indicates higher concentrations in people with HIV and green shading indicates higher concentrations in people without HIV, with darker shading for significance at a p-value < 0.05. (C) Circos plot showing correlations between cytokines measured in plasma on the left side (grey outer circle) and cytokines measured in sputum on the right side (orange outer circle). Measures from people without HIV are on the top half (purple inner circle), and from those with HIV are on the bottom half (green inner circle). Positive correlations are shown in red and negative correlations are shown in blue, with darker shading for significance at a p-value < 0.05.

With hierarchical clustering, the heatmap also uncovered distinct and heterogeneous plasma and sputum profiles with clustering of participants according to relative plasma and sputum concentrations (Figure 1B). This analysis grouped patients with HIV together as having both higher sputum and plasma concentrations. Participants with HIV also tended to have positive correlations between cytokine concentrations in sputum and plasma, whereas correlations tended to be negative in those without HIV (Figure 1C). The four cytokines with the most divergent correlations by HIV status were eotaxin/CCL11, GCSF, IL-4, and VEGF.

## Discussion

In this study of people with and without HIV who were newly diagnosed with pulmonary TB, those with HIV had significantly higher markers of inflammation in both plasma and sputum than those without HIV. As all participants had at least moderate involvement on CXR, these differences in inflammation were present despite a similar extent of radiographic involvement, suggesting that patients with TB-HIV coinfection may be particularly at risk for excessive lung inflammation. Furthermore, while correlations between plasma and sputum were generally poor, the strength and direction of correlations between blood and lung compartments differed by HIV status and people with HIV had more positive correlations than those without HIV.

Prior studies have also found that cytokine concentrations in lung samples differed from those in blood (4, 5). In addition, several studies have compared aspects of the host immune response by HIV status, including one study where people with TB-HIV coinfection had lower sputum concentrations of several matrix metalloproteinases and another where CD4 responses to *Mtb* were impaired among people with HIV (7, 8). However, these studies either did not report or did not control for the extent of radiographic involvement, which is associated with the degree of inflammation in TB.

Our finding that, among those with pulmonary TB, correlations between sputum and plasma cytokine levels were generally positive in people with HIV coinfection but negative in people without HIV may reflect impaired integrity of the lung epithelium and a propensity towards greater inflammation with HIV (1, 9). Together, these immunologic differences may increase the potential for lung damage in people with TB-HIV coinfection. If confirmed, anti-inflammatory therapies that reduce excessive lung inflammation may be particularly promising in people with TB-HIV, as found in a previous randomized trial (10).

In summary, we found that adults with pulmonary TB disease and HIV coinfection had higher levels of inflammation in both blood and lung compartments compared to those without HIV. Future studies can further explore these differences in inflammation by HIV status across the blood and lung compartments and seek to establish how these profiles may be associated with long-term outcomes and lung health after completion of TB treatment.

## Data Availability

All data produced in the present study are available upon reasonable request to the authors

## Disclaimer

The funders had no role in the study’s design, collection, analysis, or interpretation of data, the writing of the report, or the decision to submit for publication.

## Acknowledgments

We gratefully acknowledge the staff at the Aurum Institute Tembisa Clinical Research Center, including Ms. Lindelwa Nkangane, in addition to Drs. Joshua Chandler and Robert Wallis for their scientific review and input.

